# Long-term consequences of drug usage on the gut microbiome

**DOI:** 10.1101/2024.07.17.24310548

**Authors:** Oliver Aasmets, Nele Taba, Kertu Liis Krigul, Reidar Andreson, Estonian Biobank Research Team, Elin Org

**Author notes:** Collaborators Estonian Biobank research team: Mait Metspalu, Andres Metspalu, Lili Milani, Tõnu Esko. Corresponding author: Oliver Aasmets & Elin Org Institute of Genomics, Estonian Genome Centre University of Tartu Tartu 51010, Estonia.

## Abstract

Medication usage is a significant contributor to the inter-individual variability in the gut microbiome. However, drugs are often used long-term and repeatedly, a notion yet unaccounted for in microbiome studies, which might lead to underestimating the extent of drug effects. Recently, we and others showed that not only the usage of antibiotics and antidepressants at the time of sampling but also past consumption is associated with the gut microbiome. This effect can be “additive” - the more a drug is used, the stronger the effect on the microbiome. Here, by utilizing electronic health records and the Estonian Microbiome cohort metagenomics dataset (n=2,509), we systematically evaluate the long-term effects of antibiotics and human-targeted medications on the gut microbiome. We show that the past usage of medications is associated with the gut microbiome, and for example, the effects of antibiotics, psycholeptics, antidepressants, proton pump inhibitors, and beta-blockers are detectable several years after usage. Furthermore, by analyzing a subcohort (n=328) with microbiome measured repeatedly, we show that similar changes in the gut microbiome occur after treatment initiation, possibly indicating causal effects.

## Introduction

The human gut microbiome is acknowledged as an important contributor to our well-being and is considered a novel therapeutic target for health interventions. The structure and composition of this complex ecosystem reflect our health status, consumption of drugs, dietary choices, lifestyle, and the environment we live in^1–5^. As a result, a significant proportion of research is focusing on figuring out how we can use this information for disease diagnostics^6,7^, informing disease risks^8,9^ and personalization of drug usage^10,11^. However, recent evidence shows that past exposures long-preceding the sample collection can affect the gut microbiome, a direction less studied.

We have recently shown that the usage of antibiotics in the past (>6 months ago) can affect the microbiome composition independent of the antibiotic usage within 6 months of sample collection^1,12^. Moreover, this effect can be “additive” - the more drugs used in the past, the stronger the effect on the microbiome composition^1,13^. Importantly, we also showed in mice models how this long-term effect of antibiotics usage can disrupt mucus function, including mucus growth and penetrability, and may increase abdominal fat weight^14^. In addition to antibiotics, there are implications for the long-term effect of antidepressants^1^ and beta-blockers^15^, and it has been shown that the longer duration of proton pump inhibitors (PPI) consumption is associated with microbiome diversity in infants^16^. Therefore, the long-term effect of drug exposure may have a major influence on our physiology, highlighting the need to understand the full extent of such effects across diverse drug classes. However, to date, a systematic evaluation of long-term drug effects on the fecal microbiome has not been carried out.

Here, we take advantage of the possibility of linking the Estonian Microbiome cohort (EstMB) gut metagenomics data (n=2,509) with the electronic health records, which allows us to evaluate the long-term effects of drug usage systematically. We analyze the presence of long-term drug effects over a wide range of drug classes, including both human-targeted drugs (i.e. therapeutic targets of human origin) and antibiotics (i.e. therapeutic targets of bacterial origin), and assess whether the effects can be additive, as seen for antibiotics usage. Additionally, including a second time-point metagenomics data from an EstMb subcohort of 328 individuals allows us to further validate the long-term effects and study drug initiation effects.

## Results

### Drug usage in the Estonian Microbiome cohort

The Estonian Microbiome cohort (EstMB) is a population-based volunteer cohort currently including 2509 subjects (age range: 23–89 years, mean: 50.1 ± 14.93 years) who have provided blood, buccal swabs, and stool samples. In this study, we focus on the gut microbiome characterized using shotgun metagenomics sequencing. As part of the nationwide Estonian Biobank (EstBB), EstMB is supported by linkings to various electronic health records (EHR), and participants have provided data regarding their lifestyle and dietary preferences via questionnaires. A detailed overview of the EstMB cohort and available data is discussed in Aasmets & Krigul *et al.*^1^. Most importantly, EHR allows us to characterize the participants’ drug usage at the time of microbiome sampling and analyze the history of drug usage in great detail (**Fig. 1a**). Additionally, a subcohort of the EstMB (n=328) has provided a second stool sample after a median follow-up period of 4.4 years. Hereon, we refer to the first time-point of the microbiome sampling as T1 and the second time-point as T2 (**Fig. 1a**).

**Figure 1.**
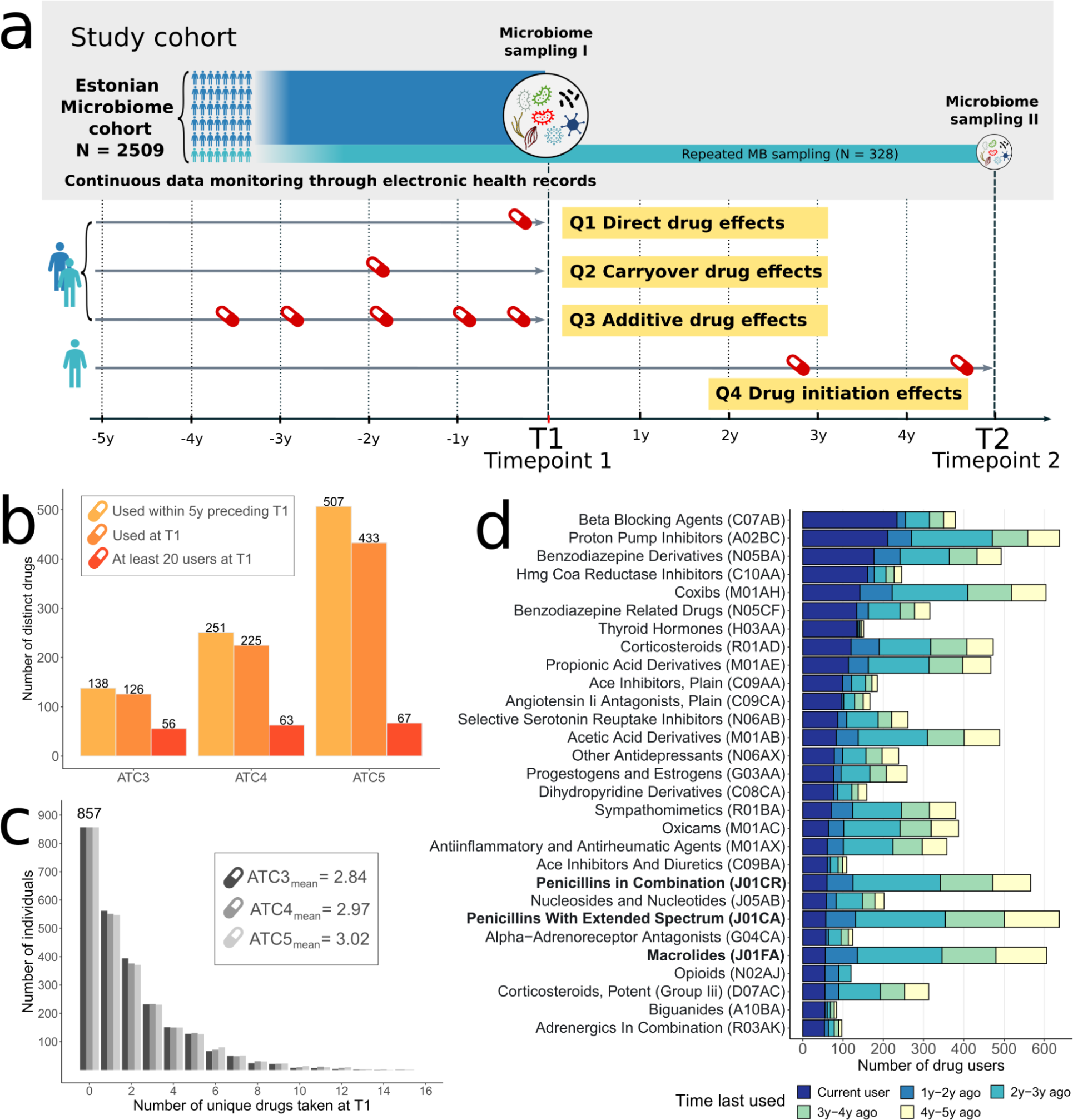
Drug usage in the EstMB cohort. **a -** Overview of the EstMB cohort and phenotype data available through the electronic health records (EHR). We aimed to analyze the direct effects of drug usage at the time of sampling (**Q1**), carryover effects induced by past drug usage (**Q2**), and additive effects, which arise from the differing amounts of drugs used in the past (**Q3**). In a subcohort of the EstMB (n=328) that has provided an additional stool sample (T2), we analyzed the alterations in the microbiome attributable to treatment initiation (**Q4**). **b -** total number of different drugs used in the EstMB at different ATC levels. **c -** distribution of the number of drugs used by the participants at T1 at different ATC levels. **d -** the number of drug users at T1 on ATC4-level; drugs with at least 50 users at T1 are shown. Drugs are ordered according to the number of active drug users. Antibiotics are highlighted in bold. ATC - Anatomical Therapeutic Chemical classification; Q - study question.

At T1, 433 prescription drugs at the ATC5 level (Anatomical Therapeutic Chemical classification system) were used by the participants, representing 225 different chemical drug subgroups at the ATC4 level and 126 pharmacological subgroups at the ATC3 level (**Fig. 1b**). Moreover, during the 5 years preceding T1, a total of 507 different medications at the ATC5, i.e. chemical substance level were used (251 at the ATC4 level and 138 at the ATC 3 level), highlighting the wide spectrum of drug usage in the population. At T1, 857 subjects (34.2%) did not use any prescription drugs, while the ones taking drugs used on average around 3 different medications from diverse drug classes at the time of microbiome sampling (ATC3 mean = 2.84; ATC4 mean = 2.97; ATC5 mean = 3.02) (**Fig. 1c**). In the downstream analyses, we focused on drugs that were used by at least 20 subjects at T1, resulting in 56 drugs at the ATC3 level, 63 drugs at the ATC4 level and 67 drugs at the ATC5 level. As an example, drugs at the ATC4 level with the most users at T1 include beta-blocking agents (n = 234, 9.3%), proton pump inhibitors (n = 211, 8.4%), and benzodiazepine derivatives (n = 177, 7.1%) (**Fig. 1d, Extended Data Table 1**), which were all often used in combination with other drugs (**Extended Data Fig. 1**). Medications with the highest number of users over the 5 years include drugs that are commonly prescribed PRN (*pro re nata*, i.e. as needed), such as proton pump inhibitors (A02BC), antibiotics (J01) and psycholeptics (N05) (**Fig. 1d**). For these drugs, a considerable number of subjects last used the drug years ago, allowing us to study long-term drug effects (**Fig. 1d**).

By taking advantage of the detailed drug usage data recorded in the EHR, we aimed to analyze the direct effects of drug usage at the time of sampling (**Q1**), drug carryover effects induced by past drug usage (**Q2**), additive effects, which arise from the different amounts of drug used in the past (**Q3**), and validate the cross-sectional findings by analyzing the effects of drug initiation between the two time-points (**Q4**) (**Fig. 1a)**.

### Effects of active drug usage on the gut microbiome composition (Q1)

Firstly, we conducted a thorough analysis to identify drugs and drug classes associated with the gut microbiome at T1. Out of the 186 drugs assessed, 167 (89.8%) were associated with either alpha diversity, beta diversity, or the abundance of at least one bacterial species (FDR <= 0.1) (**Fig. 2, Extended Data Table 2, Extended Data Table 3**). The drug effect on the beta diversity was evident in 96 of the 186 drugs (PERMANOVA on Aitchison distance; FDR <= 0.1, **Fig. 2a, Extended Data Table 2**), indicating that drugs, including human-targeted medications, can significantly alter the microbiome on the compositional scale. For example, at the chemical subgroup level (ATC4), drugs explaining the most inter-individual variability included beta-blockers (R^2^ = 0.149%, FDR = 0.0010), macrolides (R^2^ = 0.142%, FDR = 0.0010), and biguanides (R^2^ = 0.116%, FDR = 0.0010) (**Fig. 2a**, **Extended Data Table 2**). Most of these drug classes, including beta-blockers, macrolides, and benzodiazepine derivatives, are also negatively correlated with alpha diversity metrics, especially with observed richness (**Fig. 2b**). Additionally, we observe that the more unique drug classes used at the time of sampling, the lower the microbial alpha diversity (**Extended Data Fig. 2a).** Altogether, these observations are concordant with previous research and support the notion of drug usage as a significant factor in explaining inter-individual microbiome variability^5,13,17,18^.

**Figure 2.**
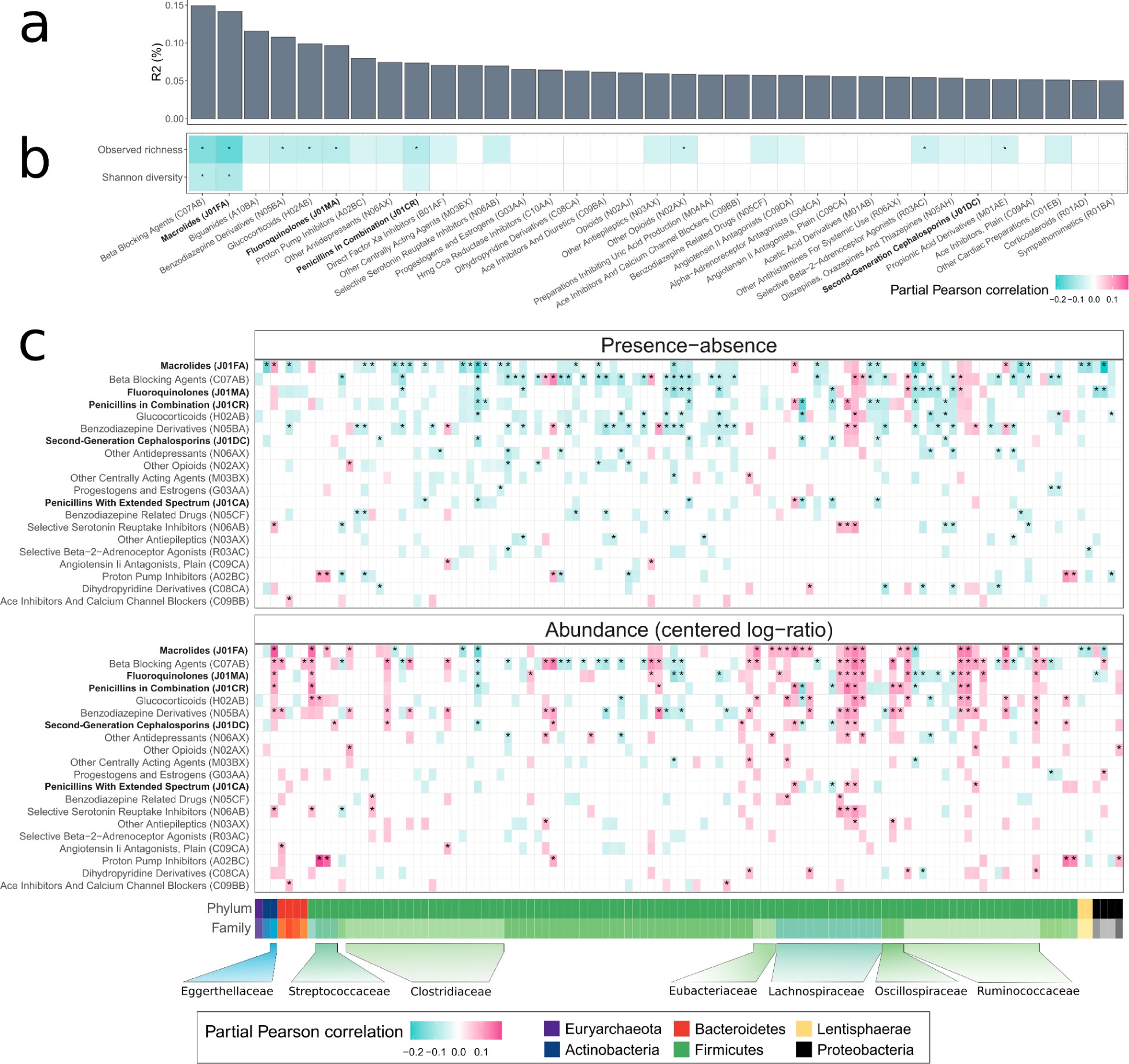
Active drug usage effects on the microbiome. (a) The interindividual variance of the microbiome explained by individual drug classes. (b) Partial Pearson correlation between drug classes at the ATC4 level and observed richness and Shannon diversity index. (c) Partial Pearson correlation between drug classes at the ATC4 level and bacterial species analyzed as presence/absence and as abundance (centered log-ratio transformed) data. The results of both analyses refer to the same bacteria, i.e., the same column refers to the same bacterial species in presence-absence and in abundance analyses. Further, only drugs and microbes with at least 10 confidently deconfounded nominally significant associations (p < 0.05) in two analyses combined are shown, while drugs are presented in descending order based on the number of associations detected (top 20 drugs shown). The colored cells indicate nominally significant and confidently deconfounded associations (p < 0.05). * - FDR <= 0.1 and confidently deconfounded. Antibiotics are highlighted in bold.

We further focused on the associations between drug usage and the presence-absence (PA) and abundance (centered log-ratio (CLR) transformed) of 530 mOTUs^19^, which were prevalent in at least 10% of the cohort. To eliminate the potential carryover effects, we compared the active drug users with the subjects who had not taken the drug of interest during the 5 years preceding T1. All analyses were adjusted for body mass index (BMI), gender, and age. To account for the potential confounding by diseases, lifestyle, and usage of other drugs, we conducted a thorough analysis, as described by Forslund *et al.*^13^. Interestingly, we identified only a few drug-bug associations that were confounded by the presence of a disease, and despite a few exceptions, we did not find significant confounding by other drugs, although such results have been shown *in vitro*^20^ (**Extended Data Table 3**). In all downstream analyses, we focused on confidently deconfounded associations. Interestingly, we found the effect directions for microbial taxa to be highly similar across drug classes, indicating a common signal of drug usage (**Fig. 2c**, **Extended Data Table 3**). This observation was evident in both abundance and presence-absence analyses. For example, *[Clostridium] asparagiforme/lavalense [06317]*, *[Clostridium] clostridioforme/bolteae [03442]* and *[Clostridium] citroniae [04828]* from the family *Lachnospiraceae* are positively correlated with the usage of beta-blocking agents, macrolides, biguanides, and PPIs, among other drugs (**Fig. 2c, Extended Data Table 3**). As human-targeted drugs have been shown to inhibit bacterial concentrations similarly to antibiotics *in vitro*^20–22^, we next examined whether a similar observation can be made *in vivo*. For that, we built machine learning models on the CLR-transformed microbiome data to predict the usage of various antibiotic subclasses and tested their ability to predict the usage of other drug classes in unobserved data. Indeed, we observed that, for example, the model aimed to detect the usage of macrolides (AUC = 0.94 for macrolides) could identify the usage of biguanides (AUC = 0.71) and selective serotonin reuptake inhibitors (AUC = 0.67), and the model aimed to detect penicillin usage (AUC = 0.75 for penicillin) can, among others, detect the usage of antidepressants (AUC > 0.58) and various corticosteroids (**Extended Data Fig. 2b, Extended Data Table 4**). In contrast, we found several bacterial species, such as *Dorea longicatena [03693]* and *Eubacterium species [12260]*, that are significantly associated with antibiotics but not with host-targeted drugs. Thus, at least partially, the effect of antibiotics and human-targeted drugs overlaps.

We then compared our results to the previously published population studies^5,13^ and the most comprehensive *in vitro* study^22^. We observed a moderate overlap with the results from population studies we could compare, especially with antibiotic and PPI usage (**Extended Data Table 5**). For example, PPIs have a positive effect on the abundance of oral microbes *Streptococcus parasanguinis* and *Veillonella parvula* in our study, in Vila *et al.*^18^, Nagata *et al.*^5^ and in Forslund *et al.*^13^. More specifically, PPIs esomeprazole and omeprazole are positively associated with *Streptococcus parasanguinis*, and esomeprazole additionally with *Veillonella parvula*. However, there were also inconsistencies - among others, for example, we observed positive associations between benzodiazepine derivatives and *Dorea formicigenerans* and *Ruminococcus torques*, whereas Nagata *et al.*^5^ observed significant negative effects for the same comparisons. Therefore, although identifying robust signals *in vivo* remains a challenge, large population-based cohorts can provide valuable insights.

We next assessed whether the drugs belonging to the same pharmacological subgroup act similarly in terms of the effect on the microbiome. We observed several notable examples with varying effects. For example, beta-blockers metoprolol (R^2^ = 0.104%; the number of univariate hits, n_univariate_ = 104) and nebivolol (R^2^ = 0.069%; n_univariate_ = 18) display remarkable differences, indicating that drugs used for a similar condition and belonging to the same pharmacological and chemical subgroup can have a different impact on the fecal microbiome. A similar discrepancy can be seen for benzodiazepine derivatives diazepam vs alprazolam and PPIs omeprazole vs pantoprazole/esomeprazole (**Extended Data Tables 2-3**). Drug dosage is a likely factor, which can complicate dissecting the bug-drug associations and be a reason for the differences in effect size for drugs belonging to the same drug group. For example, we found that the effects of PPI omeprazole on some microbes can be observed only for higher doses (**Extended Data Fig. 2c, Extended Data Table 6**). Therefore, well-powered studies that are fit to analyze the effects of distinct drugs at the chemical substance level (i.e., ATC5) and that include dosage information are likely to further elucidate the drug-bug associations.

### Long-term carryover effects on the microbiome composition are independent of recent drug usage (Q2)

Next, we focused on the identified associations with active drug usage and analyzed whether these effects were observable when the drugs were last used years before the microbiome sampling. For that, we compared the former (>1 year before T1) drug users with the subjects who had not used the drug in the 5 years preceding T1 (**Fig. 3a**, **Extended Data Fig. 3a, Extended Data Table 7**). Indeed, we observed potential carryover effects for various drug classes (78/186, 41.9% of drugs). The effects of past usage at the ATC4 level were most clearly evident for different antibiotics subclasses but also for human-targeted drugs such as benzodiazepine derivatives, biguanides, proton pump inhibitors, and antidepressants. As with active drug usage (**Fig. 2a**), the carryover effects with microbes were similar across the drug classes (**Extended Data Table 7**). Remarkably, drug-bug associations for several broad-spectrum antibiotics, such as macrolides and penicillins in combination, as well as for human-targeted drug classes, e.g. benzodiazepine derivatives and antidepressants, could be identified even if they were last used more than 3 years before the microbiome sampling (**Fig 3a**). Considering the carryover effects of antibiotics, we tried to pinpoint the duration of the effect of antibiotic usage on the microbiome richness. We observed that the diversity of antibiotic users does not seem to reach the observed richness of the antibiotic non-users, irrespective of the antibiotic load and time from the last antibiotic treatment (**Fig**. **3b**).

**Figure 3.**
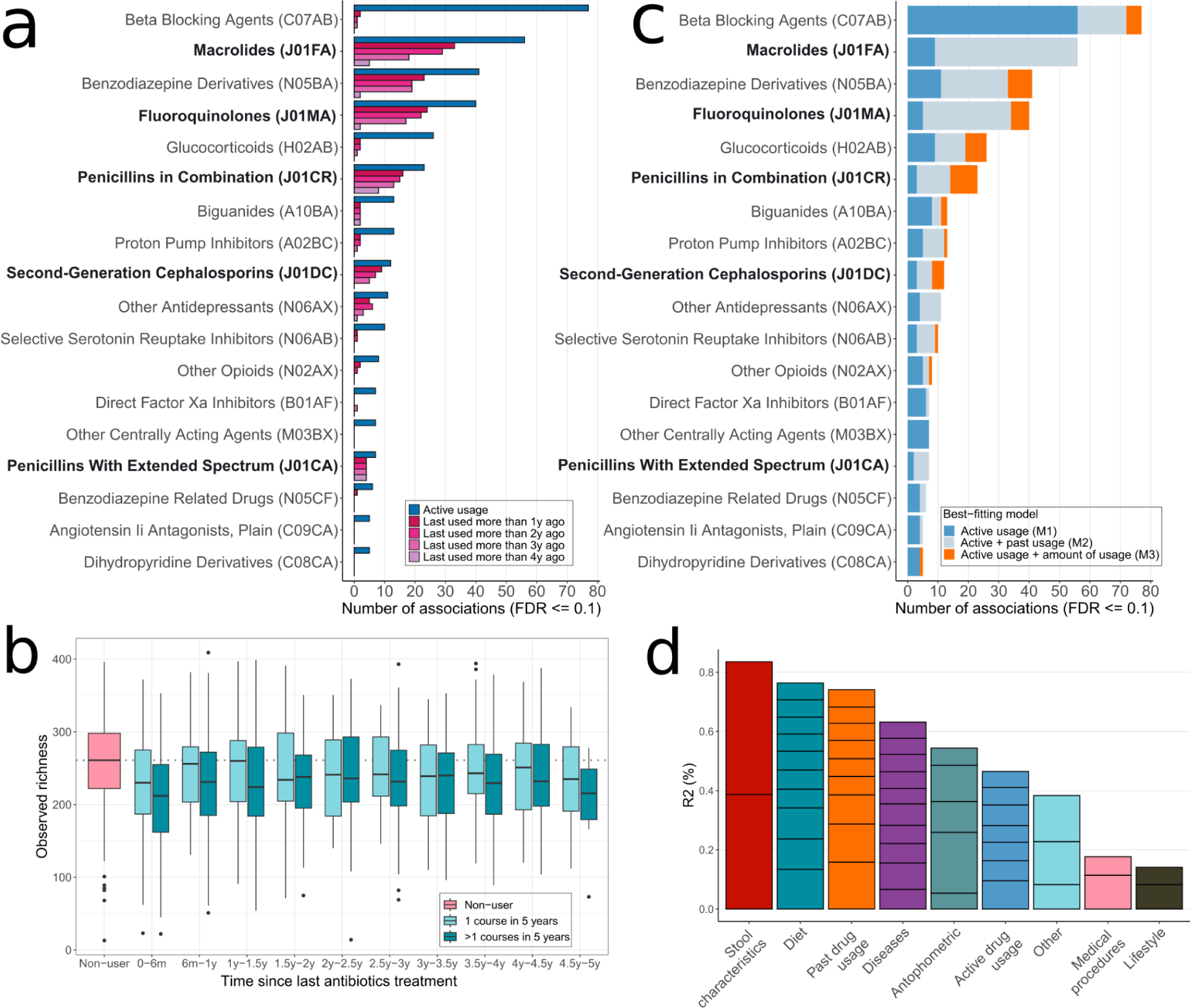
Long-term drug effects on the gut microbiome. (a) Drug carryover effects. The number of univariate associations between subjects not taking the drug and subjects having taken the drug more than 1 to 4 years prior to the microbiome sample collection (CLR-transformed abundance). (b) Long-term effect of antibiotics use on the microbiome. (c) Additive drug effects. The proportion of the univariate associations identified with active drug usage according to the model that best describes the association (CLR-transformed abundance). In addition to active drug usage as a binary trait, active drug usage together with past drug usage as a binary trait or the amount of drug usage as a continuous trait are considered. (d) Variance explained in the gut microbiome composition according to different factor groups. The bars indicating the variance explained by factor groups are further divided into individual factors according to their contribution. Past drug usage indicates drugs used during the 5-years preceding sample collection at T1. Antibiotics are highlighted in bold.

### Long-term drug effects are partly of additive nature (Q3)

The detected carryover effects raised a follow-up question, whether the long-term effects can also accumulate as previously shown for antibiotics^1,13^. Namely, does the previous drug usage or the amount of it explain additional variability in the microbiome on top of the active drug usage? To answer this question, we fit and compared three competing models (M1, M2, M3) with increasing complexity: M1 includes only active drug usage, M2 includes active and past usage (within the 5 years preceding T1) as a binary trait, and M3 includes active usage and amount of past usage measured by the number of prescriptions bought out during the 5 years preceding T1. We observed that for a majority of the drugs, the model including past usage (M2) was best-fitting for at least some of the drug-bug associations supporting the presence of long-term drug effects (**Fig. 3c**, **Extended Data Fig. 3b, Extended Data Table 8**). Moreover, the model including the amount of drug used (M3), was best-fitting for several drugs, including human-targeted drugs such as beta-blockers, benzodiazepine derivatives and glucocorticoids. For example, benzodiazepine derivatives show an additive effect on *Eisenbergiella tayi [03446]* and several *Clostridiales* species (**Extended Data Table 8**). Of note, the number of prescriptions for human-targeted drugs with continuous treatment regimes is closely related to drug adherence - the more prescriptions taken in the past, the better adherence to drug intake. Further, to characterize the effect of active and long-term drug usage on the overall variability of the fecal microbiome, we carried out a multivariate analysis of the explained variance. This analysis confirmed that long-term drug usage has a significant effect on microbiome variability, independent of active drug usage (**Fig. 3d, Extended Data Table 9**). Moreover, the long-term effects exceed the effects of active drug usage in terms of variance explained (0.74% and 0.47%, respectively). Importantly, we observe that such long-term drug usage effects can also confound disease-microbe associations, highlighting the importance of accounting for past drug usage (**Extended Data Fig. 4).**

### Microbiome measurements from the two time-points confirm long-term drug effects (Q4)

Next, we focused on the microbiome changes between T1 and T2 (median follow-up period 4.4 years) in a subcohort with the microbiome measured in two time-points (N=328). First, we compared the individuals who initiated drug usage between T1 and T2 and were active users at T2 with the drug-naive participants (**Extended Data Table 10, Extended Data Table 11**). Neither the controls nor drug initiators used the drugs 5 years before T1. Despite a limited sample size, we identified changes in the microbiome attributable to the use of drugs at T2 for broad-spectrum antibiotics, such as penicillins and macrolides, and human-targeted drugs, including proton pump inhibitors, benzodiazepine derivatives, and glucocorticoids (**Fig. 4**, **Extended Data Table 11**). We also observed drug initiation effects for drugs with no effects identified for active usage in the cross-sectional setting (Q1) (**Extended Data Table 11**). However, the proportion of nominal hits for the Q4 analysis was significantly higher in the group that also had an association with active usage in Q1 (20.5% vs 6.4%), indicating that Q1 and Q4 analyses capture similar signals. Similarly, to study the carryover effects, we compared the drug initiators, who last used the drug more than 1 year before T2, with the drug-naive subjects. This allowed us to verify the presence of carryover effects for macrolides, penicillins and human-targeted drugs such as PPIs and selective serotonin reuptake inhibitors (**Fig. 4**, **Extended Data Table 11**). All of the significant associations shown in Figure 4 were also seen in cross-sectional analyses (T1) assessing active usage (Q1) and carryover effects (Q2), with one exception: the carryover effect of macrolides (J01FA) on *Bilophila wadsworthia [04300]* was not identified. Further, among penicillins, both the active and past usage of amoxicillin and clavulanic acid (J02CR02) was significantly associated with changes (T2-T1) in the abundance of *Flavonifractor plautii [05238]*, *Oscillibacter sp. [03341]* and *Oscillibacter species incertae sedis [13092]* - all these associations were also observable in Q1 and Q2. Thus, analyzing two time-points provides a means for identifying robust drug-microbiome associations. As a last step, we analyzed the drug discontinuation between the two timepoints. The observed effects with opposite direction compared to drug initiation further support our findings.

**Figure 4.**
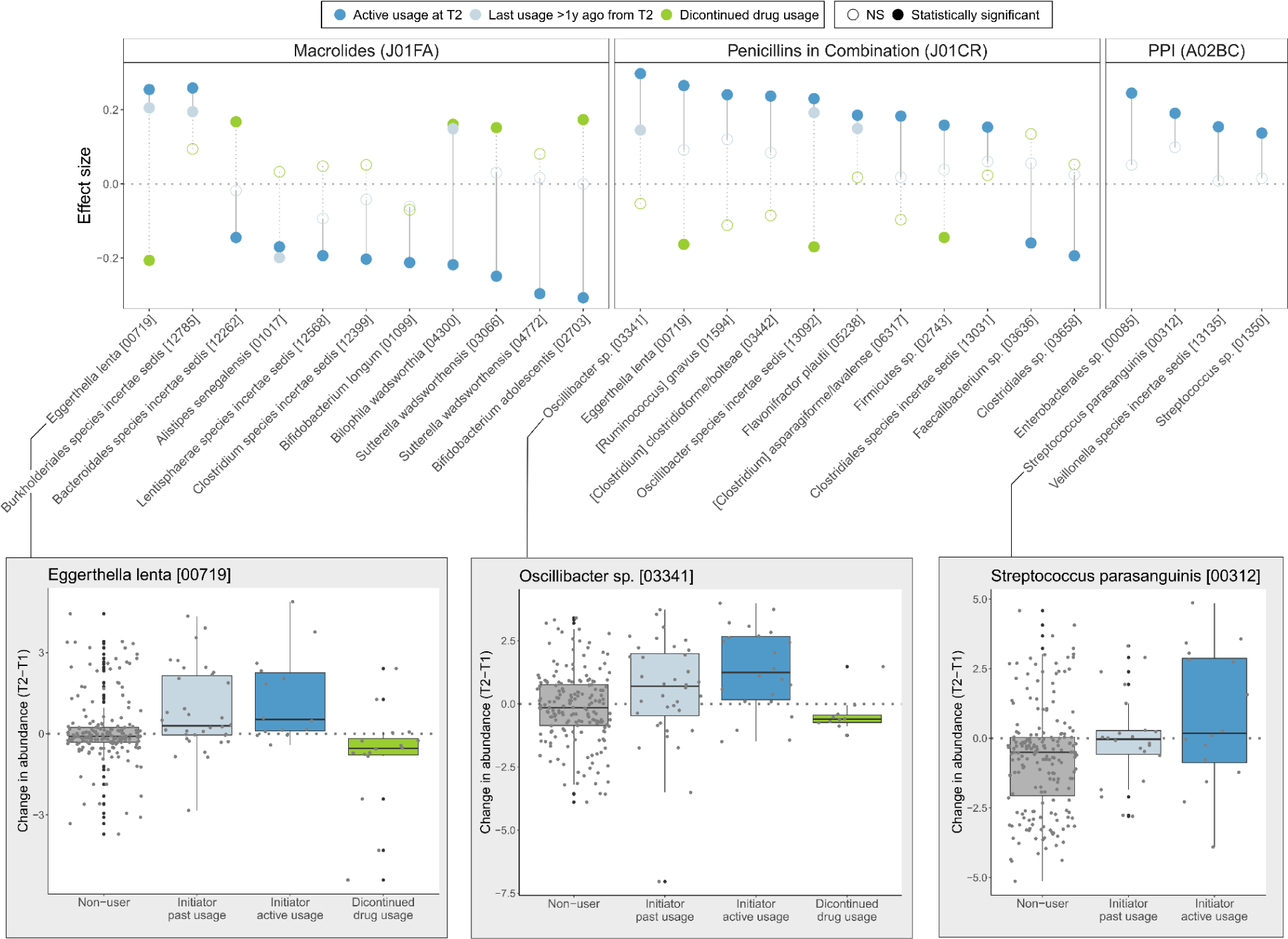
Drug initiation and discontinuation effects. Associations between drug initiation, discontinuation, and changes in the microbiome between T2 and T1 are shown for macrolides, penicillins and PPIs. Dark blue refers to associations with active drug usage, light blue refers to associations with drug usage that happened more than 1y before T2, and green refers to associations with discontinued drug use. Effect size refers to partial Pearson correlation adjusted for BMI, gender and age.

## Discussion

In this study, we carried out the first systematic evaluation of long-term drug usage effects on the fecal microbiome across various drug classes, including a large number of human-targeted drugs. While the research so far has mostly been focusing on the effects of drug use at the time of microbiome sampling, our unique dataset allowed us to demonstrate that the effect on the microbiome can still be seen years after the last usage of a drug. Taking advantage of the drug prescription data from the electronic health records available in the Estonian Biobank, we characterized the effects of drugs used within 5 years before microbiome collection. After thoroughly accounting for confounding, we observed that out of the 186 drugs analyzed, 167 (89.8%) are associated with the microbiome, while 78 of them (46.7%) display long-term effects. In addition to antibiotics, e.g. macrolides, fluoroquinolones, and different penicillin classes, several human-targeted drugs such as beta-blockers, benzodiazepine derivatives, glucocorticoids, PPIs, biguanides, and antidepressants display effects on the microbiome observable years after previous drug intake. Furthermore, this effect can be additive, i.e. it depends on the amount of drug used in the past (measured by number of prescriptions for a drug). By using second time-point samples from 328 individuals, we further verified the long-term effects of antibiotics, such as penicillins in combination and macrolides, as well as human-targeted drugs, such as PPIs and selective serotonin reuptake inhibitors, displaying a likely causal effect of these medications.

Drug usage has been previously shown to explain a significant proportion of inter-individual variability in the fecal microbiome composition^3,5,17^. Moreover, the effect of drug usage has been shown to supersede the effect of disease, further highlighting the significant impact of drug usage^5,13^. Our results extend this observation, demonstrating that past drug usage can explain additional variability independent of active drug usage, suggesting that the effect of drug usage has been underestimated. Importantly, not only antibiotics but also human-targeted drugs are among the drugs explaining additional variability. Considering that the drug burden can be highly variable across populations and specific drugs, e.g. antidepressants, antibiotics, and benzodiazepine derivatives, may exhibit varying prescription patterns across different countries^23–25^, drug usage can possibly lead to differences in the microbiome composition between the populations. For example, it has been shown that per capita antibiotic use correlates with the prevalence of antimicrobial resistance genes^26^. As antibiotic resistance mechanisms have also been shown to protect against human-targeted drugs^22^, this concurrently may affect the abundance of specific microbes in different populations and result in an overall shift in the microbiome composition. Also, the drug burden is usually higher in older age groups, so it can be expected that the microbiome in the older populations is even more affected. Similarly, clinical cohorts might also have a higher drug burden compared to the general population cohorts from volunteer-based biobanks, where healthier individuals are usually recruited. Thus, the long-term effects and varying drug burden can induce differences between cohorts and could consequently confound cross-cohort comparisons when unaccounted for. Our findings are in agreement with the current notion that several human-targeted drugs seem to act similarly to antibiotics^22^. The associations between human-targeted drugs and presence-absence of the bacteria suggest that drug usage is associated with the probability of observing bacteria in the sample. Given that most of the associations were negative, human-targeted drugs indeed seem to lower the diversity by eliminating specific bugs. The same holds for the long-term effects of human-targeted drugs. This can be one of the mechanisms that result in the long-term effects tyswe are observing. The long-term effects leave a question of whether the human-targeted drugs might also lead to detrimental microbiota-mediated physiological effects, as has been shown with the history of antibiotic use^14^. Given that human-targeted drugs are often taken continuously throughout life, not for short periods, which is the case for antibiotics, the physiological effects can be even more profound. Moreover, participants usually consume several different classes of drugs, and only a few individuals did not take any prescription medications in the 5-year observational period. Such drug-drug interactions may also be additive, i.e. long-term consumption of multiple drugs with a similar scope can supersede the detrimental effect of both individual drugs, but this remains to be studied in larger samples. Further, we identified that several disease-microbiome associations can be confounded by long-term drug usage. Therefore, disentangling disease-drug effects can be further improved by accounting for long-term drug usage in addition to active drug usage^27^ and other host variables^28^.

Surprisingly, our results showed that benzodiazepine derivatives have an even broader effect on the whole microbiome composition as well as on the presence and abundance of individual microbial species compared to several antibiotic classes. Moreover, benzodiazepine derivatives show remarkable carryover effects several years after their use, which is comparable to effects observed with the broad-spectrum antibiotics classes. The drugs belonging to benzodiazepine derivatives are well-known anti-anxiety medications that are often misused and have a high potential for drug abuse^29^. Concerningly, the use of these drugs has increased over time^30,31^. In addition, we observed that at the chemical substance level, alprazolam (ATC5 N05BA12, e.g., Xanax) and diazepam (ATC5 N05BA01, e.g. Valium) affect the microbiome in different scales, with alprazolam showing a broader impact on the microbiome. This was also previously observed by Nagata *et al.*^5^. Taking into account the rising popularity of benzodiazepines, the noted difference in the effects on the microbiome by alprazolam versus diazepam might be a valuable input for future therapy decisions and warrants further investigation. Further, the same notion could hold for other drugs, where drugs assigned for the same health condition can have an unequal magnitude of effects on the microbiome, and consequently, choosing the drug with less long-term harm on the microbiome might be favored.

*In vitro* studies have indicated that more than 100 antimicrobials and over 200 human-targeted drugs can inhibit the growth of gut commensals in isolation, which, however, might be attenuated in a community^20–22^. Nevertheless, to date, imitating the complexity of the gut microbiome and the intestinal environment remains a limitation for *in vitro* studies. Therefore, population-based cohorts using metagenomics sequencing data in combination with detailed drug usage characterization are essential to understanding the drug effects on the microbiome. One of the biggest strengths of our study is the possibility of using electronic health records (EHR) to analyze drug intake. When compared to self-reported medication data, EHR does not suffer from underreporting the use of some drugs and allows to characterize long-term drug usage. The Estonian Microbiome Cohort is a population-based volunteer cohort which allows us to study drug usage in a general population. Thus, when compared to the disease cohorts and clinical cohorts, the drug burden, general health and polypharmacy are likely to have a smaller impact. Additionally, the comprehensive statistical approach taken was supplemented by the measurements from a second time-point, which provided an internal validation of the results. We observed that more than 500 different ATC5-level drugs have been used within the last 5 years in our sample (N= 2509). However, the sample size for analyzing the effects of all of these drugs on the gut microbiome is limited. Still, comparing the results from our study to previous findings has allowed us to pinpoint several robust drug-bug interactions, such as the case of PPI usage and increase in the oral microbes, such as *S. parasanguinis* or *Veillonella parvula.* Some limitations of our study should be noted when interpreting the results. Our study focuses only on prescription-based drugs, and thus, the long-term effects of over-the-counter drugs remain to be studied. Also, we assume that when a prescription is bought out, subjects also take the drug. Similarly to other volunteer-based biobanks, our cohort suffers from gender imbalance—there are more females than males, and the cohort participants are likely to be more interested in their health.

In conclusion, our results demonstrate that medications, not limited to antibiotics, have a long-term effect on the fecal microbiome. Further, we highlight the importance of accounting for the history of drug usage when assessing disease-microbiome associations. Taken together, our results expand the understanding of drug effects on the microbiome, and we encourage researchers to focus on the long-term drug effects whenever feasible.

## Methods

### Estonian microbiome cohort and metadata preprocessing

The Estonian microbiome cohort (EstMB) was established in 2017 when stool, oral, and blood samples were collected from 2,509 Estonian Biobank (EstBB) participants. A detailed overview of the EstMB, including omics and phenotypic data availability, is described in Aasmets & Krigul *et al.* 2022^1^. Out of the 2,509 participants, 328 provided an additional stool sample after a median follow-up period of 4.4 years. We refer to the first time-point of the microbiome sampling as T1 and the second time-point as T2. All participants of the EstMB cohort gave informed consent for the data and samples to be used for scientific purposes. The study was approved by the Ethics Committee of the University of Tartu (No 266-T10) and by the Estonian Committee on Bioethics and Human Research (Estonian Ministry of Social Affairs) (No 1.1-12/2768).

Here, we rely on the Electronic Health Records (EHR) data, which is available to all the EstBB participants, including the EstMB participants. The EHR data on the diseases and medications were obtained from the Estonian Health Insurance Fund, the Estonian Cancer Registry, and the two biggest hospitals in Estonia (University of Tartu Clinic and North Estonia Medical Centre). The Anatomical Therapeutic Chemical classification (ATC) system was used to define drug classes, and drugs with at least 20 active users during the first microbiome sampling point were included. For antibiotics and antiinfectives, active usage was defined as usage within 90 days before the sample collection. For human-targeted drugs, we assessed the active drug usage according to the amount and time of purchase. This resulted in 56 drugs at the ATC3 level, 63 drugs at the ATC4 level, and 67 drugs at the ATC5 level for downstream analysis. When analyzing additive drug effects, the number of prescriptions for past usage is based on drugs purchased during the 5 years preceding the microbiome sampling, whereby the most recent prescriptions that indicate active usage are excluded from the count. We assume that when a prescription has been bought out, subjects have also consumed the drug. The summary of the number of drug users at T1 and the average number of prescriptions in 5 years per drug are summarized in **Extended Data Table 1**.

### Microbiome sample collection and DNA extraction

The participants collected a fresh stool sample immediately after defecation with a sterile Pasteur pipette and placed it inside a polypropylene conical 15 ml tube. The participants delivered the sample to the study centre, where it was stored at -80°C until DNA extraction. Microbial DNA extraction was performed using the QIAamp DNA Stool Mini Kit (Qiagen, Germany). Around 200 mg of stool was used as a starting material following the DNA extraction kit manufacturer’s instructions for the extraction. DNA was quantified from all samples using a Qubit 2.0 Fluorometer with dsDNA Assay Kit (Thermo Fisher Scientific). NEBNext® Ultra™ DNA Library Prep Kit for Illumina (NEB, USA) was used for generating sequencing libraries following the manufacturer’s recommendations. Briefly, 1 μg DNA per sample was used as input material. Index codes were added to attribute sequences to each sample. The DNA sample was fragmented by sonication to an average size of 350 bp, DNA fragments were end-polished, A-tailed, and ligated with the full-length adaptor for Illumina sequencing with further PCR amplification. Finally, PCR products were purified (AMPure XP system), and libraries were analyzed for size distribution using Agilent2100 Bioanalyzer and quantified using real-time PCR.

### Metagenomics data analyses

The shotgun metagenomic paired-end sequencing was performed by Novogene Bioinformatics Technology Co., Ltd. using Illumina NovaSeq6000 platform, resulting in 4.62 ± 0.44 Gb of data per sample (insert size 350 bp, read length 2 x 250 bp). First, the reads were trimmed for quality and adapter sequences. The host reads that aligned to the human genome were removed using *SOAP2.21* (parameters: -s 135 -l 30 -v 7 -m 200 -x 400)^32^. The taxonomic profiling was done using the *mOTUs2.5* tool with default parameters^19^. In total, 14 213 marker gene-based operational taxonomic units (mOTUs) were identified. Alpha and beta-diversity analyses were carried out on the whole identified composition. For univariate analysis, mOTUs, which were detected in at least 10% of the samples, were used to limit the number of tests carried out, resulting in 530 mOTUs. Filtered mOTU profiles were also used as predictors for building classification models. We did not rarefy the counts to avoid loss of data.

### Statistical analysis

All statistical analyses were done using the R (v. 4.0.1) software.

### Diversity analysis

We used observed richness and the Shannon diversity index to assess alpha diversity. Shannon index was calculated using the *vegan* package (v2.5-6)^33^. Associations between observed richness, Shannon index, and drug usage were analyzed as described in the univariate analysis section. To calculate the between-sample distances for beta diversity analysis, we used the Euclidean distance on the centered log-ratio (CLR) transformed microbiome species-level profile^34^. We tested the associations between drug usage and microbiome composition with Permutational Analysis of Variance (PERMANOVA^35^) on the between-sample distances using 10,000 permutations for the p-value calculation (**Extended Data Table 2)**. To carry out PERMANOVA, we used the *adonis* function from the *vegan* package. To apply the CLR transformation, zero-counts were imputed with a pseudocount equal to half of the minimal non-zero relative abundance value.

### Univariate analysis

To analyze active drug usage effects (Q1 in **Fig. 1a**), we compared subjects taking a drug at T1 with subjects who had not taken the drug during the 5 years preceding T1. To analyze the effect of human-targeted drugs, we excluded subjects who had used antibiotics within the 90 days preceding T1. To associate the abundance of each mOTU with the drug usage, we used linear models adjusted for age, BMI, and gender:

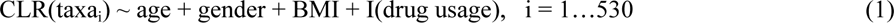

We used the same linear regression models for observed richness and Shannon diversity index. We report the partial Pearson correlation coefficient that is adjusted for age, BMI, and gender as the effect size. Alternatively, to associate the presence and absence (PA) of each mOTU as a binary trait with the drug usage, we used logistic regression models adjusted for age, BMI, and gender:

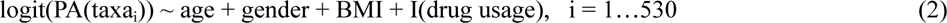

where we considered relative abundance value >0 to indicate the presence of a taxa. We accounted for multiple testing using the Benjamini–Hochberg procedure.

To analyze the carryover effects (Q2 in **Fig. 1a**), we compared subjects who had last taken the drug more than x years (x = 1, 2, 3, 4) before T1 with those who had not taken the drug during the 5 years preceding T1. Similarly to active drug usage, linear and logistic regression models, as in formulas (1) and (2), were used for the analysis. For carryover analysis and other downstream analyses, we compared the bug-drug pairs where the effect of active drug usage was identified (analysis Q1, FDR <= 0.1) (**Extended Data Table 7**).

To analyze additive drug effects (Q3 in **Fig. 1a**), we compared the fit of three competing models:

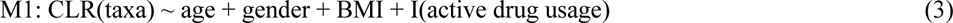

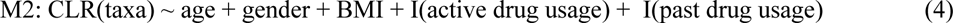

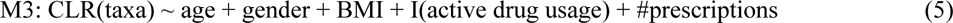

Models are increasing in complexity: Model M1 indicates only the effect of active drug usage, M2 indicates the effect of active and past usage, and M3 indicates the effect of active drug usage and the amount of drug usage during the past 5 years as indicated by the number of prescriptions bought out in the past (independent of active usage). Akaike information criteria (AIC) was used to compare the fit of the models. A more complex model was considered to be a better fit if the difference in AIC with the simpler model was more than 2 (**Extended Data Table 8**). Additionally, we performed the same analysis with presence/absence data, where in M1, M2, and M3, instead of CLR(taxa) is logit(PA(taxa)).

To study the drug initiation effects (Q4 in **Fig. 1a**), we compared the subjects initiating the drug usage between T1 and T2 with the subjects who did not use the drug between the timepoints or at T2. Additionally, neither the controls nor drug initiators used the drugs 5 years before T1. We further divided the drug initiators into drug initiators who used drugs at T2 and initiators who last used the drug more than a year before T2 to study the active drug usage effects and carryover drug effects. Drugs with at least 10 initiators or discontinuers were analyzed (**Extended Data Table 10**). We used linear models adjusted for age, BMI, and gender to analyze drug initiation-related changes in the abundances of each mOTU between T1 and T2 (**Extended Data Table 11)**:

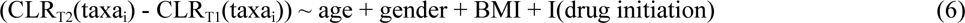

Additionally, we compared the subjects who were using the drug at T1 but did not use the drug between T2 and T1 with the subjects who hadn’t used the drug at all to study the effects of drug discontinuation (Q4 in **Fig. 1a**).

To analyze the effect of different drug dosages, we used linear models adjusted for age, BMI, and gender (**Extended Data Table 6)**:

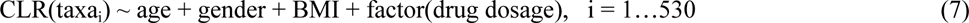

A likelihood ratio test was carried out to test the significance of drug dosage in the model.

### Deconfounding analysis

We carried out a rigorous *post hoc* analysis for Q1 and Q2 to identify potential confounding factors for the bug-drug associations as described by Forslund *et al.*^13^. Firstly, we identified naive associations between bugs and drugs, as described in the previous section. Next, for each covariate considered as a potential confounder, we fit a pair of nested linear models adjusted for BMI, age, gender, and covariate to assess whether a) the predictive ability of the drug exceeds the predictive ability of the covariate or b) the predictive ability of the covariate exceeds the predictive ability of the drug. The association identified in the first step was considered confounded when, for at least one covariate, the covariate’s predictive ability exceeded the drug’s predictive ability, but the opposite was not true. All prevalent diseases, all other drugs, lifestyle, anthropometric, and dietary factors described in Aasmets & Krigul *et al.*^1^ were considered in the analysis as potential confounding factors. The deconfounding analysis was similarly applied to identify confounders for disease-microbiome interactions. For that, the number of prescriptions for all of the drugs was considered as potential confounders in addition to the aforementioned factors to identify disease-microbiome interactions that are confounded by past drug usage. Naive disease-drug associations were identified using the linear regression model adjusted for gender, BMI, and age at sampling.

### Prediction analysis for antibiotics usage

We fit regularized linear models to predict, based on the CLR-transformed microbiome data, the active usage of antibiotic subclasses (at ATC4 level with at least 50 users at T1 (**Extended Data Table 1**)). We implemented the elastic net models in R using the *tidymodels* (v0.1.1) and *glmnet* (v3.0-2) packages. First, we split the data in a 75:25 ratio to the training and test datasets. The models for each antibiotic subclass were tuned on the training data using a 4-time repeated 5-fold cross-validation and grid search with 50 hyperparameter combinations. The initial data split and cross-validation splits were stratified by the drug usage to address class imbalance. Next, we evaluated whether the models built to detect antibiotic usage could identify the usage of other human-targeted drugs. For that, we assessed the models’ ability to predict the usage of non-antibiotic drugs in the test dataset, using the area under the receiver operating characteristic (AUROC) (**Extended Data Table 4**). For the evaluation, we excluded the antibiotic users from the test set. The model building and evaluation were repeated 5 times on random training-test splits, and the performance estimates were averaged.

### Multivariate analysis of variance components

We used a distance-based redundancy analysis to evaluate the amount of explained variance by each factor and factor group. As factors, we considered prevalent diseases, anthropometric, lifestyle, and dietary factors as described in Aasmets & Krigul *et al.*^1^, as well as active drug usage as binary traits and past drug usage indicated by the number of prescriptions bought out in the past (independent of active usage) as continuous traits. We fit the initial model by combining all the factors using the *dbrda* function from the *vegan* package, followed by a forward-selection model fitting procedure using the *ordistep* function. For each factor in the selected model, we assessed how much it can explain the community variation, accounting for all other selected variables (**Fig. 3d**, **Extended Data Table 9**). Euclidean distance on the centered log-ratio (CLR) transformed microbiome species-level profile was used to calculate between-sample distances.

## Data availability

The metagenomic data are available upon request. The phenotype data contain sensitive information from electronic health registers and they are available through the EstBB upon submission at https://genomics.ut.ee/en/biobank.ee/data-access.

## Code availability

The source code for the analyses is available at https://github.com/oliveraasmets17/EstMB_drugUsage.

## Acknowledgements

The authors thank the Estonian Biobank research team (Mait Metspalu, Andres Metspalu, Lili Milani, Tõnu Esko) from the Estonian Genome Centre, Institute of Genomics, University of Tartu, for collection of the health records data for the EstBB. Additionally, the authors would like to thank Mari-Liis Tammesoo, Marili Palover, Anu Reigo, Neeme Tõnisson, Liis Leitsalu, Triinu Temberg, and Esta Pintsaar for participating in the sample collection process. We thank Steven Smit, Rita Kreevan, and Martin Tootsi for the DNA extraction process.

## Author Contributions

EO and OA designed, and EO supervised the study. OA performed the statistical analysis. RA performed the bioinformatic analysis of shotgun metagenomics sequencing data. OA, NT and KLK interpreted the data and prepared the figures. OA, NT, KLK, RA and EO wrote the manuscript. All authors read and approved the final manuscript.

## Extended Data Tables

1. Extended Data Table 1. Number of drug users
2. Extended Data Table 2. Beta diversity analysis
3. Extended Data Table 3. Univariate analysis, active usage effects
4. Extended Data Table 4. Machine learning analysis
5. Extended Data Table 5. Univariate analysis, literature overlap
6. Extended Data Table 6. Drug dosage analysis
7. Extended Data Table 7. Univariate analysis, carryover effects
8. Extended Data Table 8. Univariate analysis, best fit (AIC)
9. Extended Data Table 9. Multivariate variance partitioning analysis
10. Extended Data Table 10. Number of drug initiators
11. Extended Data Table 11. Univariate analysis, drug initiation

## Extended Data Figures

**Extended Data Fig. 1.**
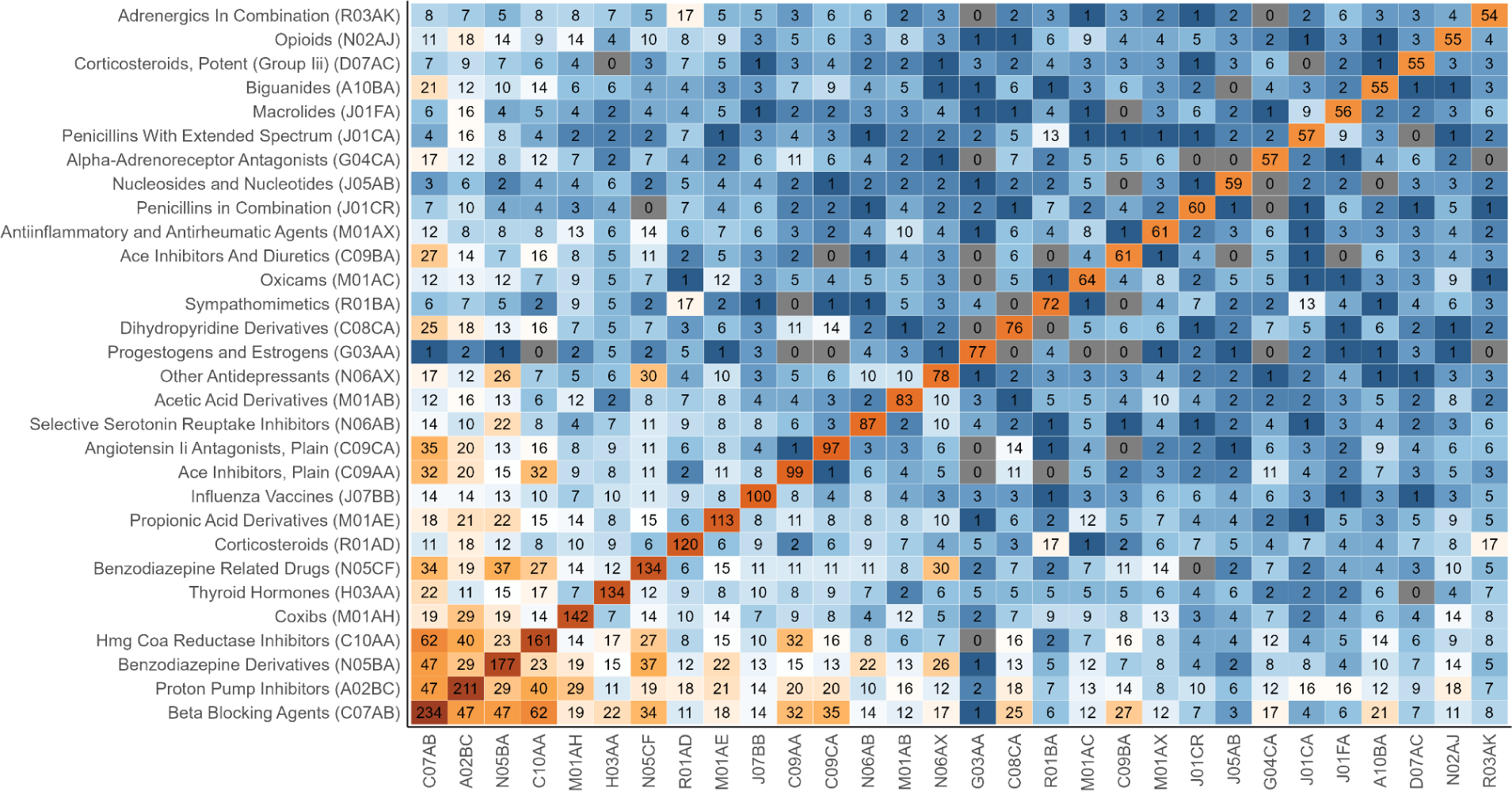
Number of participants using the drugs (ATC4-level) and drug combinations at T1. The number of participants using a specific drug is shown diagonally, and the number of subjects using the combination of the drugs is shown off-diagonally.

**Extended Data Fig. 2.**
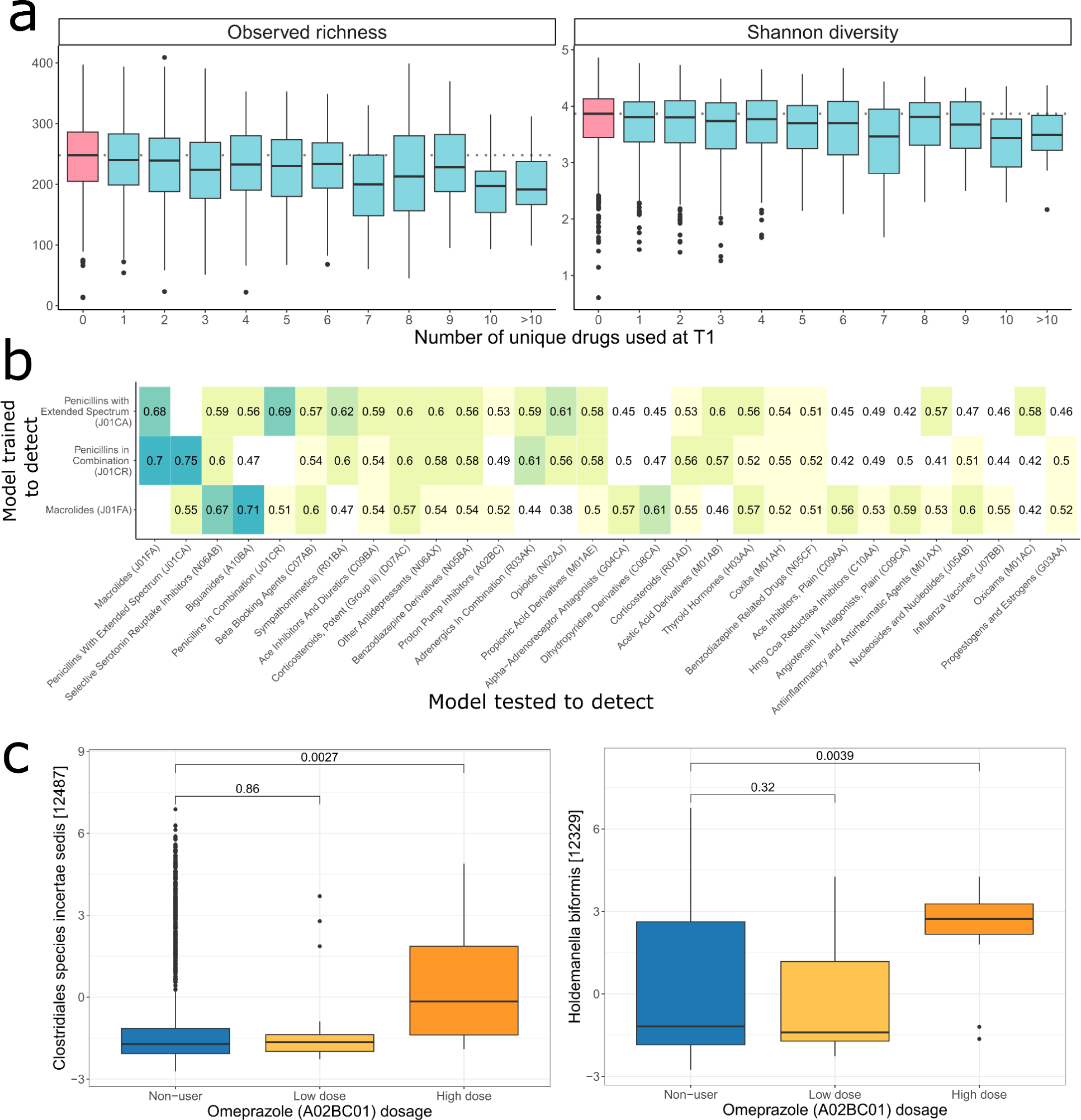
Active drug usage. (a) Alpha diversity measures (Observed richness and Shannon diversity) associated with the number of prescriptions at T1. (b) The performance of machine learning models aimed at detecting antibiotic usage is applied to detect the usage of the drugs. Values show the model’s performance (Area Under the Receiver Operating Characteristics AUROC) in an independent test set AUROC. (c) Associations between selected individual species CLR-transformed abundance and PPI omeprazole dosage. *Post-hoc* t-test p-values are shown for the group comparisons.

**Extended Data Fig. 3.**
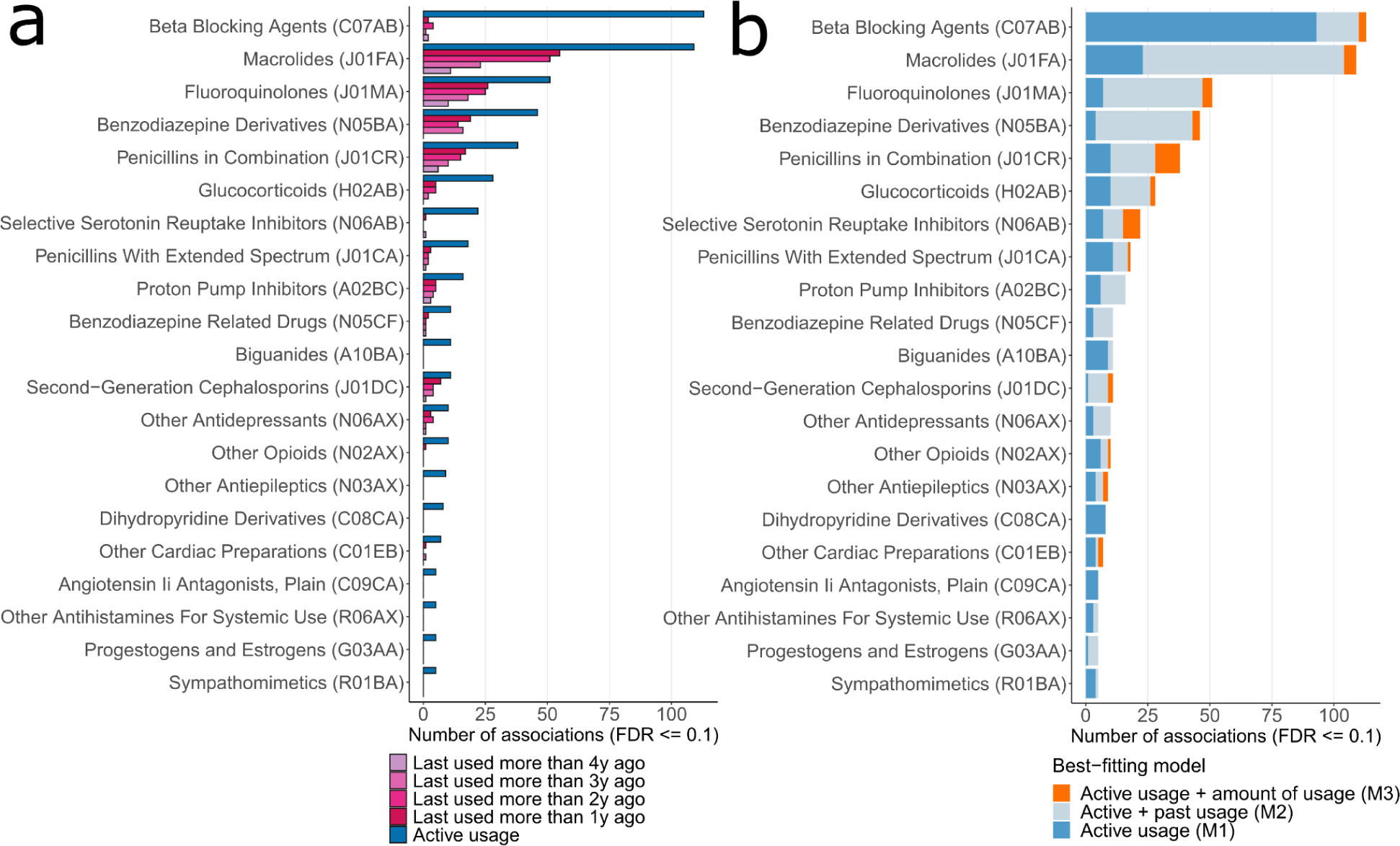
Number of univariate associations with microbiome presence-absence (PA). (a) Drug carryover effects. The number of univariate associations between subjects not taking the drug and subjects having taken the drug more than 1 to 4 years prior to the microbiome sample collection. (b) Additive drug effects. The proportion of the univariate associations identified with active drug usage according to the model that best describes the association. In addition to active drug usage as a binary trait, active drug usage together with past drug usage as a binary trait or the amount of drug usage as a continuous trait are considered.

**Extended Data Fig 4.**
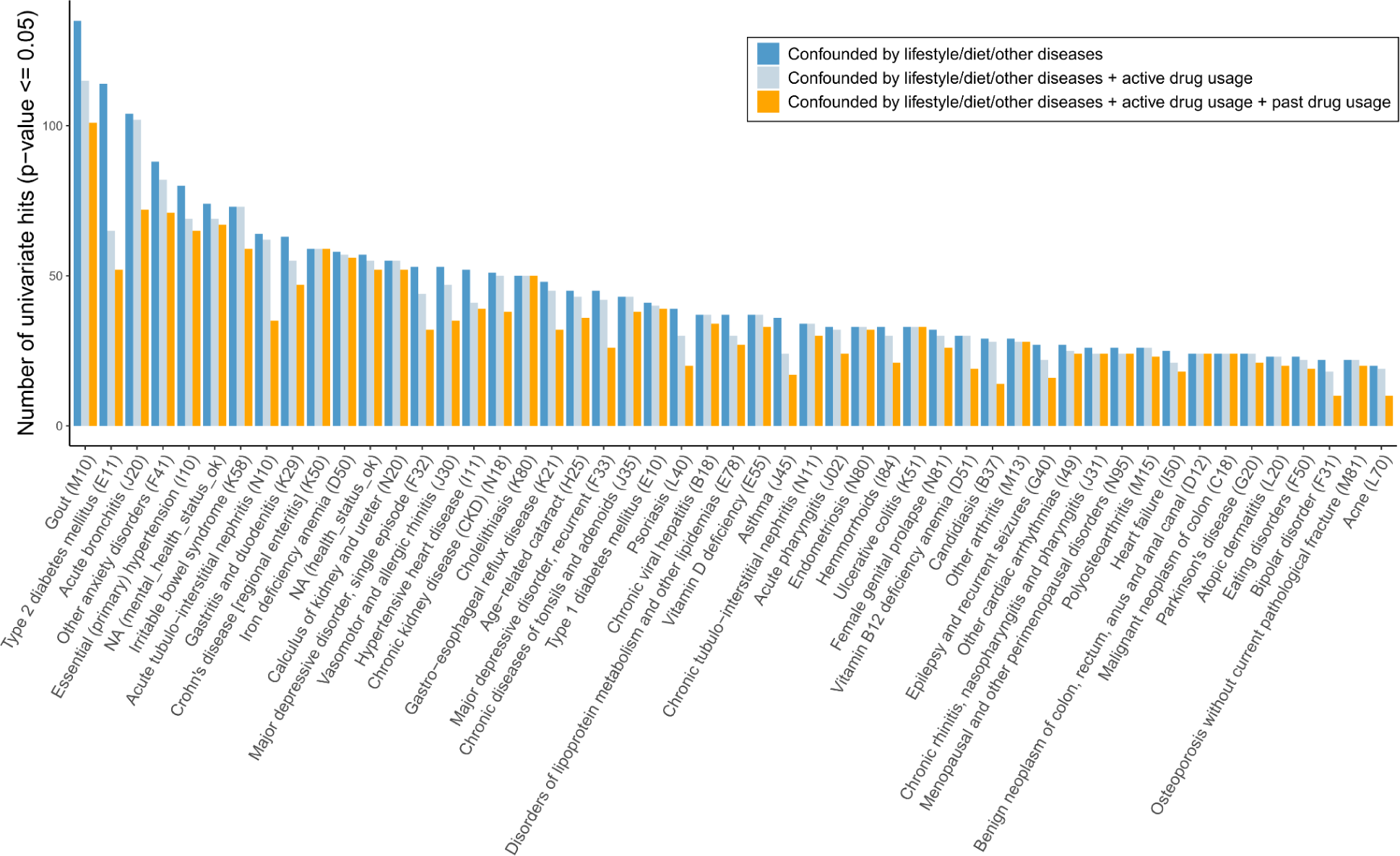
Number of univariate associations between the CLR-transformed abundance of bacterial species and prevalent diseases after deconfounding analysis.

## References

1. Aasmets, O., Krigul, K. L., Lüll, K., Metspalu, A. & Org, E. Gut metagenome associations with extensive digital health data in a volunteer-based Estonian microbiome cohort. Nat. Commun. 13, 869 (2022).

2. Falony, G. et al. Population-level analysis of gut microbiome variation. Science 352, 560–564 (2016).

3. Gacesa, R. et al. Environmental factors shaping the gut microbiome in a Dutch population. Nature 604, 732–739 (2022).

4. Zhernakova, A. et al. Population-based metagenomics analysis reveals markers for gut microbiome composition and diversity. Science 352, 565–569 (2016).

5. Nagata, N. et al. Population-level Metagenomics Uncovers Distinct Effects of Multiple Medications on the Human Gut Microbiome. Gastroenterology 163, 1038–1052 (2022).

6. Kartal, E. et al. A faecal microbiota signature with high specificity for pancreatic cancer. Gut 71, 1359–1372 (2022).

7. Wirbel, J. et al. Meta-analysis of fecal metagenomes reveals global microbial signatures that are specific for colorectal cancer. Nat. Med. 25, 679–689 (2019).

8. Liu, Y. et al. Early prediction of incident liver disease using conventional risk factors and gut-microbiome-augmented gradient boosting. Cell Metab. 34, 719–730.e4 (2022).

9. Salosensaari, A. et al. Taxonomic signatures of cause-specific mortality risk in human gut microbiome. Nat. Commun. 12, 2671 (2021).

10. Gopalakrishnan, V. et al. Gut microbiome modulates response to anti–PD-1 immunotherapy in melanoma patients. Science 359, 97–103 (2018).

11. Wilmanski, T. et al. Heterogeneity in statin responses explained by variation in the human gut microbiome. Med 3, 388–405.e6 (2022).

12. Aasmets, O., Krigul, K. L. & Org, E. Evaluating the clinical relevance of the enterotypes in the Estonian microbiome cohort. Front. Genet. 13, (2022).

13. Forslund, S. K. et al. Combinatorial, additive and dose-dependent drug–microbiome associations. Nature 600, 500–505 (2021).

14. Krigul, K. L. et al. A history of repeated antibiotic usage leads to microbiota-dependent mucus defects. 2024.03.07.583875 Preprint at 10.1101/2024.03.07.583875 (2024).

15. Si, J. et al. Long-term life history predicts current gut microbiome in a population-based cohort study. *Nat*. Aging 2, 885–895 (2022).

16. Brusselaers, N., Pereira, M., Alm, J., Engstrand, L. & Engstrand Lilja, H. Effect of proton pump inhibitors in infants with esophageal atresia on the gut microbiome: a pilot cohort. Gut Pathog. 14, 47 (2022).

17. Jackson, M. A. et al. Gut microbiota associations with common diseases and prescription medications in a population-based cohort. Nat. Commun. 9, (2018).

18. Vila, A. V. et al. Impact of commonly used drugs on the composition and metabolic function of the gut microbiota. Nat. Commun. 11, 1–11 (2020).

19. Milanese, A. et al. Microbial abundance, activity and population genomic profiling with mOTUs2. Nat. Commun. 10, 1014 (2019).

20. Brochado, A. R. et al. Species-specific activity of antibacterial drug combinations. Nature 559, 259–263 (2018).

21. Garcia-Santamarina, S. et al. Emergence of community behaviors in the gut microbiota upon drug treatment. 2023.06.13.544832 Preprint at 10.1101/2023.06.13.544832 (2023).

22. Maier, L. et al. Extensive impact of non-antibiotic drugs on human gut bacteria. Nature 555, 623–628 (2018).

23. Bruyndonckx, R. et al. Consumption of antibiotics in the community, European Union/European Economic Area, 1997–2017. J. Antimicrob. Chemother. 76, ii7–ii13 (2021).

24. Lewer, D., O’Reilly, C., Mojtabai, R. & Evans-Lacko, S. Antidepressant use in 27 European countries: associations with sociodemographic, cultural and economic factors. Br. J. Psychiatry J. Ment. Sci. 207, 221–226 (2015).

25. Lukačišinová, A. et al. Prevalence, country-specific prescribing patterns and determinants of benzodiazepine use in community-residing older adults in 7 European countries. BMC Geriatr. 24, 240 (2024).

26. Lee, K. et al. Population-level impacts of antibiotic usage on the human gut microbiome. Nat. Commun. 14, 1191 (2023).

27. Forslund, K. et al. Disentangling type 2 diabetes and metformin treatment signatures in the human gut microbiota. Nature 528, 262–266 (2015).

28. Vujkovic-Cvijin, I. et al. Host variables confound gut microbiota studies of human disease. Nature 587, 448–454 (2020).

29. Casati, A., Sedefov, R. & Pfeiffer-Gerschel, T. Misuse of medicines in the European Union: a systematic review of the literature. Eur. Addict. Res. 18, 228–245 (2012).

30. Kurvits, K., Toompere, K., Jaanson, P. & Uusküla, A. The COVID-19 pandemic and the use of benzodiazepines and benzodiazepine-related drugs in Estonia: an interrupted time-series analysis. Child Adolesc. Psychiatry Ment. Health 18, 66 (2024).

31. Maust, D. T., Lin, L. A. & Blow, F. C. Benzodiazepine Use and Misuse Among Adults in the United States. Psychiatr. Serv. 70, 97–106 (2019).

32. Li, R. et al. SOAP2: An improved ultrafast tool for short read alignment. Bioinformatics 25, 1966–1967 (2009).

33. Oksanen, J. et al. vegan: Community Ecology Package. (2024).

34. Aitchison, J. The Statistical Analysis of Compositional Data. J. R. Stat. Soc. Ser. B Methodol. (1982) doi:10.1111/j.2517-6161.1982.tb01195.x.

35. Anderson, M. J. A new method for non-parametric multivariate analysis of variance. Austral Ecol. 26, 32–46 (2001).

